# Consensus-Based Definitions for Vocal Biomarkers: The International VOCAL Initiative

**DOI:** 10.1101/2025.10.23.25338518

**Authors:** Mégane Pizzimenti, Ayush Kalia, Jamie A. Toghranegar, Mohamed Ebraheem, Nicholas Cummins, Satrajit S. Ghosh, James T. Anibal, Rhoda Au, Arian Azarang, Ruth H. Bahr, Steven D. Bedrick, Hugo Botha, Oita C. Coleman, Abir Elbeji, Lampros C. Kourtis, Anaïs Rameau, Jaskanwal Deep Singh Sara, Stephanie W. Watts, Daria Hemmerling, Jiri Mekyska, Marisha L. Speights, the Bridge2AI Voice Consortium, the eVoiceNet COST Action (CA24128), Jean-Christophe Bélisle-Pipon, Yael E. Bensoussan, Guy Fagherazzi

**Affiliations:** Deep Digital Phenotyping Research Unit, Department of Precision Health, Luxembourg Institute of Health, Strassen, Luxembourg; USF Health Voice Center, Department of Otolaryngology-Head & Neck Surgery, University of South Florida, Tampa, FL, United States; Department of Internal Medicine, Yale School of Medicine, New Haven, United States; Department of Biostatistics & Health Informatics, Institute of Psychiatry, Psychology and Neuroscience, King’s College London, London, United Kingdom; McGovern Institute for Brain Research, Massachusetts Institute of Technology, Cambridge, MA, United States; Center for Interventional Oncology, NIH Clinical Center, National Institutes of Health, Bethesda, United States; Computational Health Informatics Lab, Institute of Biomedical Engineering, University of Oxford, Oxford, United Kingdom; Department of Medicine, Boston University Chobanian & Avedisian School of Medicine, Boston, Massachusetts, United States; Department of Anatomy & Neurobiology, Boston University Chobanian & Avedisian School of Medicine, Boston, Massachusetts, United States; Boston University Alzheimer’s Disease Research Center, Boston, Massachusetts, United States; The Framingham Heart Study, Boston University Chobanian & Avedisian School of Medicine, Boston, Massachusetts, United States; Department of Epidemiology, Boston University School of Public Health, Boston, Massachusetts, United States; Department of Neurology, Boston University Chobanian & Avedisian School of Medicine, Boston, Massachusetts, United States; Department of Radiology, The University of North Carolina at Chapel Hill, Chapel Hill, NC, United States; Department of Communication Sciences & Disorders, University of South Florida, Tampa, FL, United States; Departments of Medical Informatics and Clinical Epidemiology and Medicine, Oregon Health and Science University, Portland, OR, United States; Department of Neurology, Mayo Clinic, Rochester, MN, United States; Open Voice TrustMark Initiative, Apex, NC, United States; Alzheimer’s Drug Discovery Foundation, Boston, MA, United States; Sean Parker Institute for the Voice, Department of Otolaryngology-Head & Neck Surgery, Weill Cornell Medical College, New York, NY, United States; Department of Cardiovascular Medicine, Mayo Clinic, Rochester, MN, United States; Minneapolis Heart Institute and Minneapolis Heart Institute Foundation, Abbott Northwestern Hospital, Minneapolis, Minnesota, United States; Department of Measurement and Electronics, AGH University of Krakow, Kraków, 30-059, Poland; Department of Telecommunications, Brno University of Technology, Brno, Czech Republic; Central European Institute of Technology (CEITEC), Masaryk University, Brno, Czech Republic; Department of Communication Sciences and Disorders, Northwestern University, Evanston, Illinois 60202, United States; Department of Bio-ethics, Faculty of Health Sciences, Simon Fraser University, Burnaby, British Columbia, Canada

**Keywords:** Digital Health, Digital Voice, Digital Speech, Biomarker, Digital Biomarker, Vocal Biomarker, Voice Biomarker, Speech Biomarker, AI, Signal Processing

## Abstract

**Importance:** Voice-based health technologies are growing rapidly, but they lack standardized terminology, which hinders interdisciplinary collaboration, research quality, and clinical translation.

**Objective:** The objective of this work is to develop universally accepted definitions in the rapidly evolving field of vocal biomarkers, as part of the VOCAL (<u>V</u>ocal Biomarker Guidelines for <u>O</u>ntology, <u>C</u>lassification, <u>A</u>pplication, and <u>L</u>ogistics) initiative, a structured, international consensus-based framework that aims to provide standards, and guidelines.

**Design:** VOCAL is a rigorous, international, multi-stage consensus-building study conducted in 2024-2025.

**Setting:** Multi-institutional collaboration between representatives from the Bridge2AI-Voice Consortium (North America) and the eVoiceNet Network (European Union), culminating in an in-person workshop at the 2025 Bridge2AI Voice Symposium.

**Participants:** A group of 24 international experts in medicine, clinical research, speech and language, audio signal processing, statistics, methodology, regulation, ethics.

**Methods:** VOCAL’s iterative process involved five rounds of review, feedback, and an in-person workshop at the international 2025 Bridge2AI Voice Symposium, ensuring the incorporation of diverse perspectives and achieving a robust agreement on the proposed definitions.

**Main outcomes and measures:** Consensus-based definitions for vocal biomarkers, spanning from broad concepts (biomarker, digital biomarker, vocal biomarker) to domain-specific measures (cardio-respiratory acoustic, voice, speech/articulatory, cognitive/language).

**Results:** A hierarchical continuum model of vocal biomarkers was established. We first distinguished between the concepts of vocal measures and vocal biomarkers. We then defined terms from broad, overarching concepts (Level 0: Biomarker, Digital Biomarker, Vocal Biomarker) to more specific physiological and cognitive domains (Level 1: Cardio-Respiratory Acoustic; Level 2: Voice; Level 3: Speech/Articulatory; Level 4: Cognitive/Language, including linguistic and paralinguistic subtypes).

**Conclusions and Relevance:** This work provides a shared vocabulary that is essential for fostering communication through interdisciplinary collaboration, improving the quality and efficiency of research and development, and ensuring the ethical, reliable, and scalable deployment of future voice-based health technologies. It lays foundational groundwork for upcoming guidelines and standards, which are crucial for advancing the field of vocal biomarkers into widespread clinical utility.

**Key points:** *Question:* Can we establish an international consensus on the definitions related to vocal biomarkers?

*Findings:* Through a multi-stage international consensus process involving expert representatives from the Bridge2AI-Voice Consortium and eVoiceNet Network, a hierarchical model of vocal biomarkers was developed, defining terms from broad concepts (biomarker, digital biomarker, vocal biomarker) to specific physiological and cognitive domains (cardio-respiratory acoustic, voice, speech/articulatory, cognitive/language).

*Meaning:* Standardized definitions for vocal biomarkers provide an essential shared vocabulary for interdisciplinary collaboration and lay the foundational groundwork for future guidelines needed to advance the implementation of voice-based health technologies into clinical practice and clinical research.

## Introduction

In recent years, the concept of vocal biomarkers has garnered considerable attention for its transformative potential in detecting, monitoring, and managing various health conditions through the analysis of voice-based technologies.^1,2^ Vocal biomarkers fall under the broader category of digital biomarkers, which are objective and quantifiable health indicators derived from data collected via digital devices such as wearables and sensors. These non-invasive digital biomarkers promise to revolutionize clinical trials and healthcare delivery by enabling continuous, objective, and patient-centered data collection.^3^

A key barrier to making vocal biomarkers a reality is the presumption that any digitally acquired voice metric is a vocal biomarker, despite not having undergone rigorous validation. Making a parallel with imaging and blood-based biomarkers, extensive research and investment over many years have gone into validating Alzheimer’s disease imaging and blood-based measures as biomarkers. This same careful, scientific approach for biomarker validation is not currently being applied to digital data. When validating any biomarker, the context of use must be defined to establish it as a biomarker for a specific disease. Thus, moving the field from measuring features of the voice and speech to validated vocal biomarkers will require the time, energy, and investment necessary to undergo validation processes that biological biomarkers have already undergone. The path to validated biomarkers is generally defined by authorities such as the Food and Drug Administration (FDA) or the European Medicines Agency (EMA). Guidelines for digital biomarkers from these regulatory agencies are still in their nascent stages. Thus, there is an opportunity to collaborate with them in developing validation pipelines for various types of vocal biomarkers. We provide definitions of different types of vocal biomarkers below, with the caveat that validation is needed for each subtype within each category and for a specific context of use. Future work will require a transparent validation process and address concerns regarding artificial intelligence (AI)-driven approaches to vocal biomarker identification, details of which are discussed below.

Once validated, vocal biomarkers will be unique among digital biomarkers in that they derive from the interplay of multiple physiological systems (respiratory, phonatory, articulatory, and cognitive) captured through audio. Their integration into healthcare reflects a broader movement toward more real-time and remote health sensing.^4,5^ In Parkinson’s disease, speech and language impairments are common and often precede motor dysfunction; features such as reduced loudness, monotone speech, imprecise articulation, and altered rhythm show strong potential for early detection and tracking disease severity and progression.^6^ In Alzheimer’s disease, lexical-semantic and acoustic measures show high diagnostic performance for mild cognitive impairment, correlate with amyloid-β status and hippocampal volume, and predict 2-year disease progression, supporting their use for early detection and monitoring.^7^ Psychiatric and neurodevelopmental conditions likewise exhibit distinctive vocal patterns: for example, depressed individuals tend to speak more slowly, quietly, and with flattened prosody (with frequent pauses), and these acoustic characteristics correlate with depression severity.^8^ In chronic heart failure, voice analysis offers a promising, cost-effective, and convenient alternative for diagnosing, risk prediction, and telemonitoring clinical outcomes.^9^ Vocal biomarkers also help identify smoking status from ecological audio recordings,^10^ and show promise as a screening or monitoring tool for conditions like type 2 diabetes.^11^

Despite this rapid growth, a fundamental challenge persists: the absence of universally accepted definitions for key terms such as “voice biomarkers”, “vocal biomarkers”, and “speech biomarkers”, among others. These terms are frequently used interchangeably in the literature,^10,12–18^, yet they reflect features originating from distinct physiological and cognitive systems. A review of existing research reveals a wide variance in definitions, often leading to nearly as many interpretations as there are articles on the topic.^4^ Such terminological ambiguity presents a significant barrier to scientific progress.^19^ It hampers collaboration across diverse disciplines, complicates efforts to share data, and impedes the development of standardized methodologies and best practices.^20^ Without a common language, the field risks fragmentation and inefficiency, ultimately undermining its scientific advancement, regulatory assessment, clinical translation, and trustworthiness of its innovation.^21,22^

This challenge is further reinforced by a set of interrelated risks that threaten the field’s coherence and scalability. The widespread use of AI, proprietary algorithms, and closed datasets has created a “black box” problem, where the lack of transparency limits reproducibility and erodes trust among researchers, clinicians, and regulators. In parallel, inconsistent terminologies and disciplinary silos exacerbate confusion, highlighting the urgent need for a shared, evolving vocabulary. The technical landscape adds further complexity, as voice data is often massive, heterogeneous, and collected under diverse conditions, making integration across studies difficult without adherence to FAIR (Findable, Accessible, Interoperable, Reusable) principles.^23^ Voice data-specific regulatory frameworks remain fragmented and difficult to navigate, particularly across regions, posing significant hurdles to clinical validation and implementation.^24^ Additionally, concerns about privacy, data ownership, and ethical use of sensitive voice data persist,^25^ alongside practical challenges such as ensuring sustained patient engagement in digital trials.^26^ A recent survey of stakeholders in voice AI development emphasized the need for ethically sourced, diverse, and transparent data practices, calling for trustworthiness not just as a technical feature but as a guiding norm.^25^

The lack of consensus also impairs the development of methodological infrastructure. While master protocols are now standard in clinical trials and other biomarker fields, vocal biomarker research lacks unified frameworks, resulting in heterogeneous data collection, feature extraction, and analysis methods.^4^ This absence complicates validation and clinical scalability. These converging barriers underscore the pressing need for community-driven standards, transparent methodologies, and harmonized regulatory pathways to unlock the full potential of vocal biomarkers. Establishing a unified framework for vocal biomarkers is therefore essential. Standardization will provide a shared vocabulary that facilitates interdisciplinary collaboration, supports data interoperability, and enables reproducibility of research findings. This foundation is critical for advancing vocal biomarker research from exploratory studies to reliable clinical applications.

To address this challenge, this paper proposes a set of foundational definitions as the first step of the VOCAL initiative (standing for <u>V</u>ocal Biomarker Guidelines for <u>O</u>ntology, <u>C</u>lassification, <u>A</u>pplication, and <u>L</u>ogistics), a structured, international framework for harmonizing practices and guiding the field of vocal biomarkers. We aim to develop a shared and controlled vocabulary that promotes clarity and consistency across disciplines. This will facilitate effective communication among all stakeholders and the general public, and it will guide the development of best practices for the field.

## Methods

### Consensus-Building Process

To develop clear and widely applicable definitions of vocal biomarkers, a structured, multi-stage consensus-building process was employed. This approach was meticulously designed to incorporate diverse perspectives from interdisciplinary stakeholders, ensuring transparency and rigor throughout the development of the framework. The iterative nature of the process enabled continuous refinement and validation of the proposed definitions, resulting in a robust and widely accepted outcome.

### Participants

The consensus process involved three distinct groups of participants, ensuring broad representation and a comprehensive range of expertise:

● **Internal Team (n = 7):** This group comprised core members directly involved in the initial drafting of definitions and the coordination of the entire consensus process. They were leaders (YB, GF) and representatives of two large international consortia on vocal biomarkers: the Bridge2AI Voice Consortium and the eVoiceNet network. Their expertise spanned critical areas, including laryngology, speech-language pathology, digital epidemiology, AI, research methodology, and bioethics. This diverse composition ensured a strong foundational understanding of both clinical and technical aspects of vocal biomarkers.
● **Selected Group of Experts (n = 5):** This group consisted of international academic researchers with highly specialized expertise in speech analysis, clinical research, and the development and commercialization of vocal biomarkers. Their focused input provided in-depth technical and methodological critique, refining the precision and scientific accuracy of the definitions.
● **Broader Expert Group (n = 18):** This extended panel comprised a diverse array of researchers, clinicians, voice AI researchers, and stakeholders from a wide range of relevant disciplines (Clinical and Medical Sciences, Speech and Language Sciences, Engineering and Technical Fields, Statistics and Methodology, Regulation, Ethics, and Translation). This group provided a comprehensive review and validation of the definitions from various practical and theoretical standpoints, ensuring their applicability across the broader scientific and clinical landscape.

### Iterative Process

The process was organized into five iterative rounds of review and feedback, systematically designed to progressively refine the definitions and achieve strong consensus among all participants (see Figure 1; Supp Table 1).

● **Initial Draft and First Round of Review:** The internal team initiated the process by drafting preliminary definitions of vocal biomarkers. A comprehensive review informed this of existing literature and terminology. Following this, team members individually reviewed and provided comments on the draft. A subsequent group meeting was held to discuss and consolidate all feedback, leading to the first set of revisions (Revision #1).
● **Second Round of Review:** The revised definitions (Revision #1) were then shared with both the internal team and the selected group of experts. Participants provided written feedback, which was then discussed in an online workshop. This workshop focused on identifying points of agreement and divergence, and the collective input led to a second set of revisions (Revision #2).
● **Third Round of Review:** The updated definitions (Revision #2) were again reviewed by the internal team and the selected experts. A second online workshop was convened to refine terminology further and address any remaining ambiguities or concerns. This meticulous discussion resulted in a third revision (Revision #3), which brought the definitions closer to a final consensus.
● **Fourth Round of Review:** The extensively revised definitions (Revision #3) were presented to the broader expert group for review and finalization. An in-person workshop was conducted at the Voice AI Conference in Tampa, April 2025, during which participants formally voted by a show of hands on their agreement or disagreement with each presented definition. Any definition receiving disagreement from 25% or more of the participants was subjected to further discussion and revision. This iterative refinement continued until a consensus was achieved for all definitions.
● **Fifth Round of Review and Finalization**: Feedback from the workshop was further integrated into a final set of definitions developed by the internal team. Following this final round, all co-authors reviewed and approved the definitions and the overarching framework presented in this paper.

**Figure 1.**
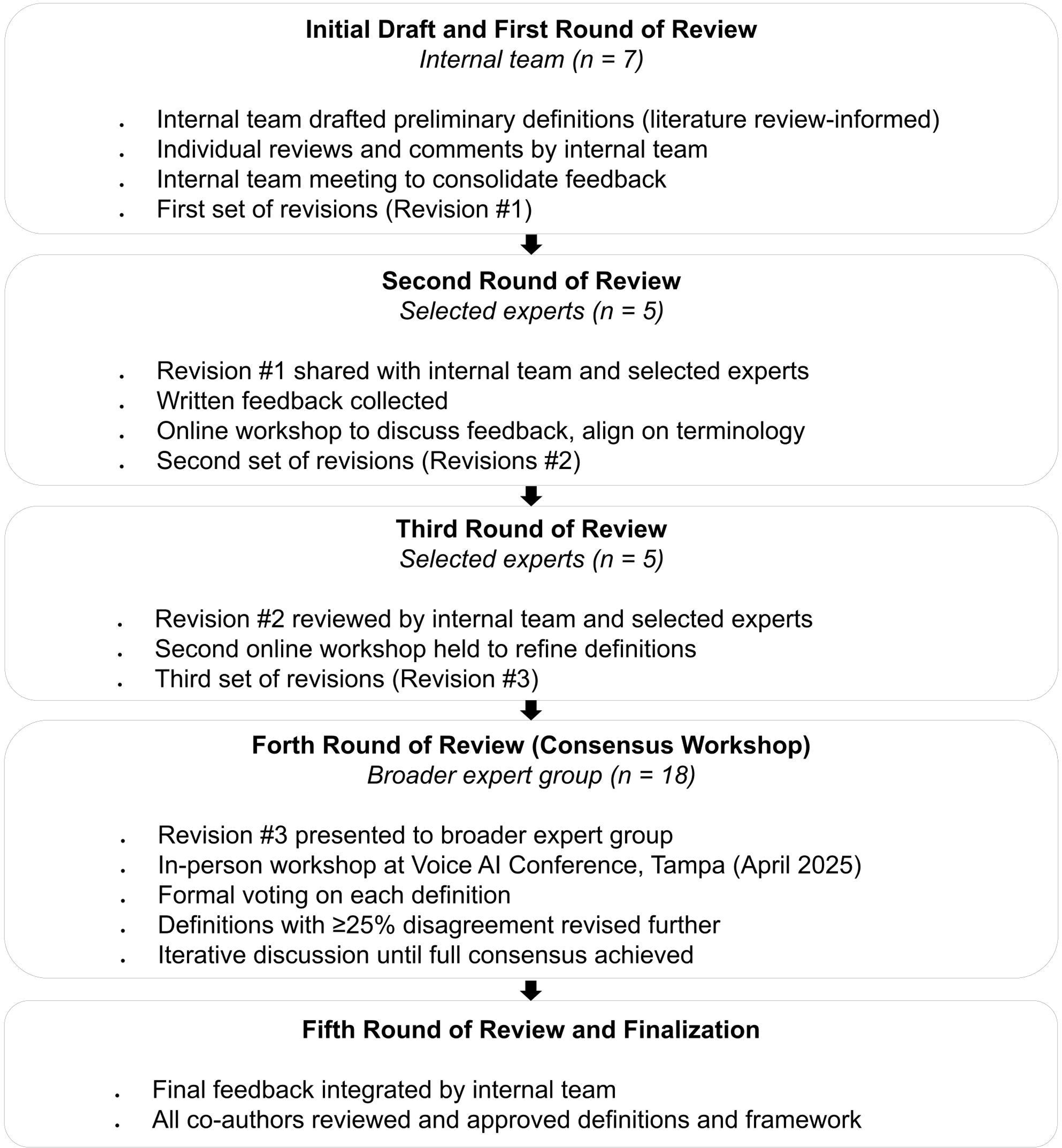
Flowchart of the iterative process to establish the definitions of key terms of the VOCAL initiative. The process included an initial internal team draft and first review, two rounds of review with selected experts, a consensus workshop with a broader expert group, and a final review and approval. The number of participants at each stage is indicated (n).

**Table 1.** Level 0 Definitions.

| Term | Definition | Explanatory Notes/Examples |
| --- | --- | --- |
| **Biomarker** | A measurable biological substance or characteristic whose detection or evaluation provides information about the function or state of a system or organ in the body. It can be used to understand normal or altered biological processes, and may serve roles in risk prediction, screening, diagnosis, monitoring, treatment response measurement, or prognosis. | There are various types of biomarkers, including physiological, serological, imaging, and genetic ones. For instance, HbA1c serves as a blood biomarker for diabetes. Another example of a biomarker is the presence of the BRCA gene, which indicates a genetic predisposition to certain types of breast cancer. In line with regulatory definitions, a biomarker is formally recognized only after undergoing appropriate validation (analytical and clinical). However, within the research and innovation context, especially in emerging areas such as voice-based health technologies, the term "biomarker" is commonly used pre-validation to denote a candidate indicator under investigation. |
| **Digital Biomarker** | A biomarker derived from data collected using a digital device such as a wearable, portable, or implantable device. | For example, a heart rate measured from a smartwatch is a digital biomarker, and an ECG measured by a wearable is a digital biomarker. A digital biomarker is a digital measure that can be used as a digital endpoint in a clinical study. However, not all digital measures are digital biomarkers. |
| **Vocal Biomarker**<br>(overarching term, includes voice, speech, linguistic, paralinguistic, and prosody) | A vocal biomarker is an overarching term for defining biomarkers derived from audio recordings of the voice, speech, linguistic, and paralinguistic | It is important to understand that these should not represent mutually exclusive categories, as they can overlap and should be interpreted more as a continuum. Moreover, |
|  | systems. These are collected through acoustic data recording and can be analyzed using traditional acoustic analysis or digital signal processing and AI methods. This overarching term includes biomarkers that are impacted by the process of creating speech from our lungs and cardio-respiratory system to our larynx, resonators, and brain, which helps with lexical access and influences the content of our speech. | extracted features can be interpreted as different types of biomarkers, depending on the context.<br>Example: F0 can be used as a voice biomarker in dysphonia or voice disorders, and can also be considered in mental health screening for depression. |

This rigorous process ensured that all concerns were thoroughly addressed through collective discussion before the definitions were finalized.

## Results

To capture the layered complexity of vocal biomarkers while maintaining clarity and usability, the definitions were structured across different levels of granularity, forming a continuum model. This hierarchical approach enables both a broad conceptual understanding and precise categorization based on the underlying physiological and cognitive systems involved in voice production.

● **Level 0** represents broad, overarching terms in the biomarker domain. These foundational definitions are intended to frame the scope of discussion and provide shared reference points across diverse disciplines and technologies.
● **Levels 1 to 4** provide more specific and structured definitions of terms that fall under the umbrella of vocal biomarkers. These are categorized based on the stage or domain of vocal production and analysis, ranging from the earliest physiological processes to higher-level cognitive functions (see Figure 2 and Figure 3).

**Figure 2.**
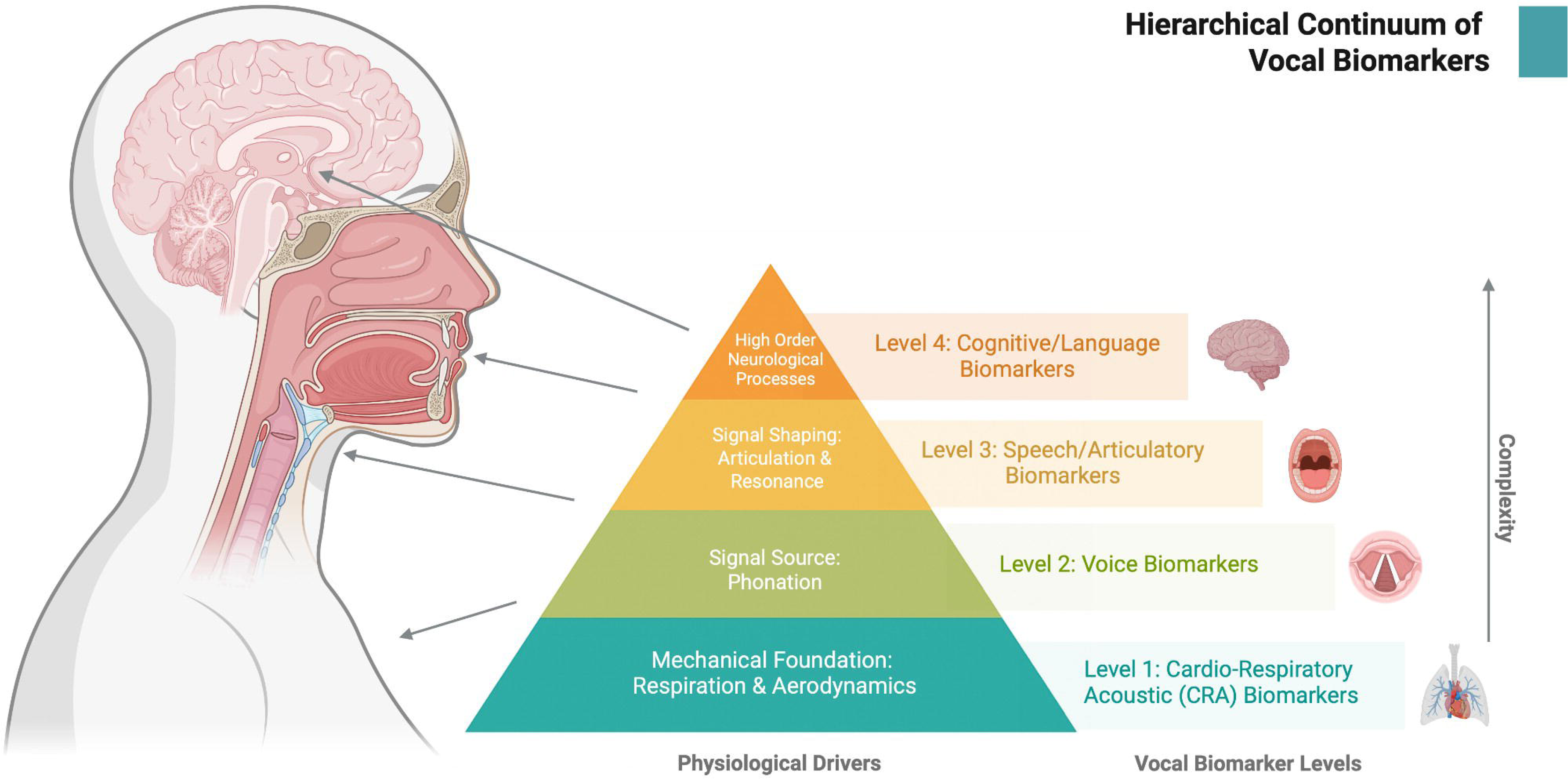
Hierarchical continuum of vocal biomarkers: from voice production to high-level cognition. The pyramid illustrates the hierarchy of vocal biomarkers, organized by physiological drivers and complexity. **Level 1** biomarkers (blue) represent cardio-respiratory acoustic biomarkers driven by respiration and aerodynamics. **Level 2** biomarkers (green) reflect voice biomarkers arising from phonation. **Level 3** biomarkers (yellow) capture speech and articulatory biomarkers shaped by articulation and resonance. **Level 4** biomarkers (orange) represent cognitive and language biomarkers arising from high-order neurological processes. Complexity increases from the base to the apex of the pyramid. Corresponding anatomical regions associated with each biomarker type are shown on the left. Created in https://BioRender.com

**Figure 3.**
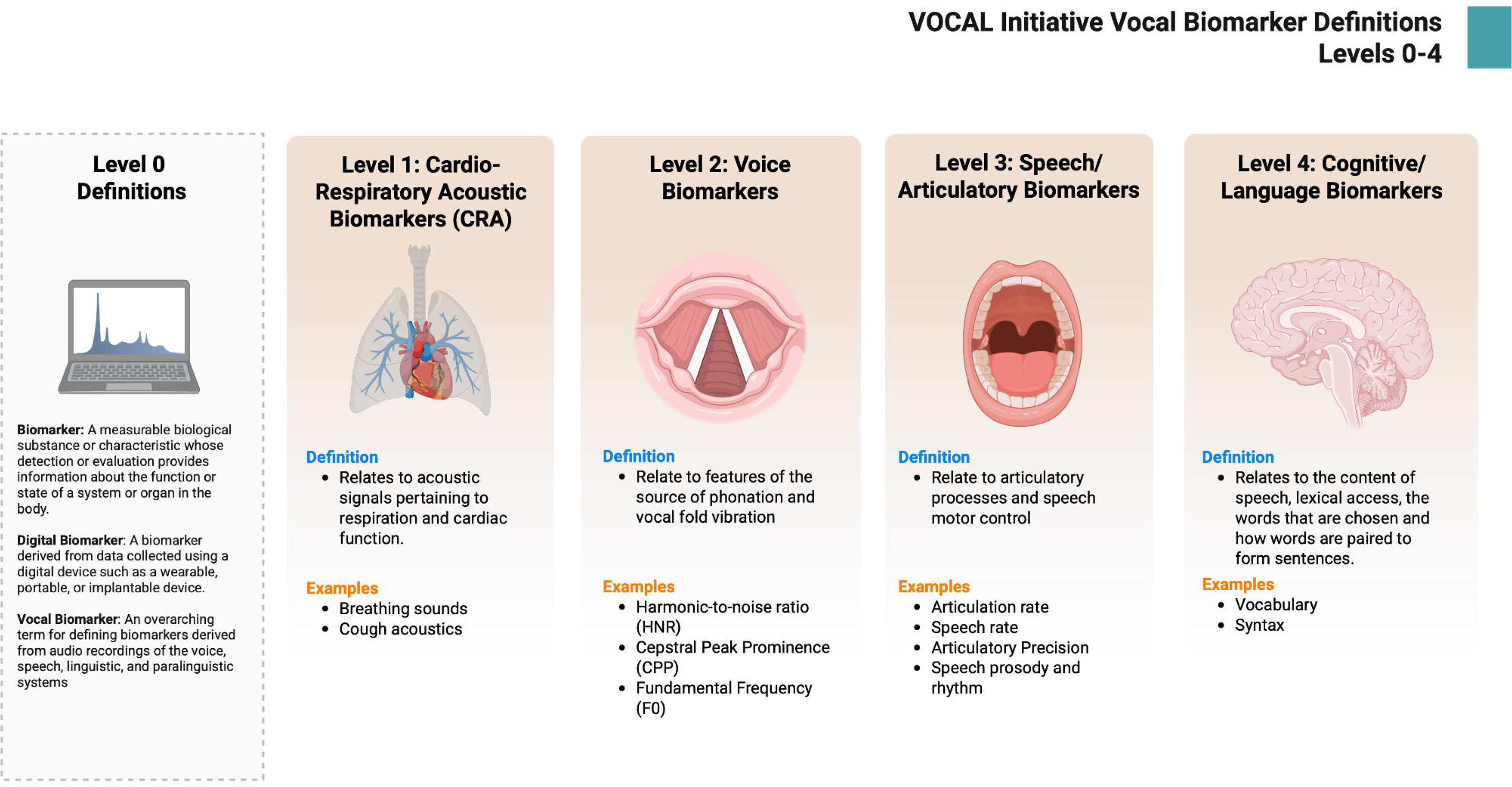
Vocal Biomarkers Levels 0-4. Vocal biomarkers can be categorized across four levels based on their physiological and cognitive origins. **Level 0: Biomarker, Digital Biomarker, Vocal biomarker** represent broad, overarching terms. **Level 1: Cardio-Respiratory Acoustic Biomarkers (CRA)** relate to acoustic signals associated with respiration and cardiac function (e.g., breathing sounds, cough acoustics). **Level 2: Voice Biomarkers** pertain to features of phonation and vocal fold vibration (e.g., harmonic-to-noise ratio, cepstral peak prominence, fundamental frequency). **Level 3: Speech/Articulatory Biomarkers** involve articulatory processes and speech motor control (e.g., articulation rate, speech rate, articulatory precision, speech prosody and rhythm). **Level 4: Cognitive/Language Biomarkers** reflect the content of speech, including lexical access, word pairing, syntax, and paralinguistic elements such as emotion, cultural cues, and dialects (e.g., vocabulary, syntax). Created in https://BioRender.com

It is crucial to understand that these categories are not mutually exclusive and often overlap, reflecting the interconnected nature of human voice production. Furthermore, extracted features can be interpreted as different types of biomarkers, depending on the specific context in which they are applied.

### Level 0 Definitions

Table 1 establishes the foundational terminology for the entire framework, ensuring that readers from diverse backgrounds, including clinicians, engineers, researchers, and regulators, have a common understanding of “Biomarker,” “Digital Biomarker,” and “Vocal Biomarker” before delving into more specific categories (see Supp Figure S1). This clarity is paramount for interdisciplinary discourse and regulatory alignment.

### Level 1-4 Definitions

Table 2 breaks down the complex domain of vocal biomarkers into actionable, physiologically grounded categories. By providing clear definitions, examples, and illustrations, it serves as a practical guide for researchers and clinicians to accurately classify and interpret vocal features, thereby reducing ambiguity and facilitating standardized research design and reporting.

**Table 2.**
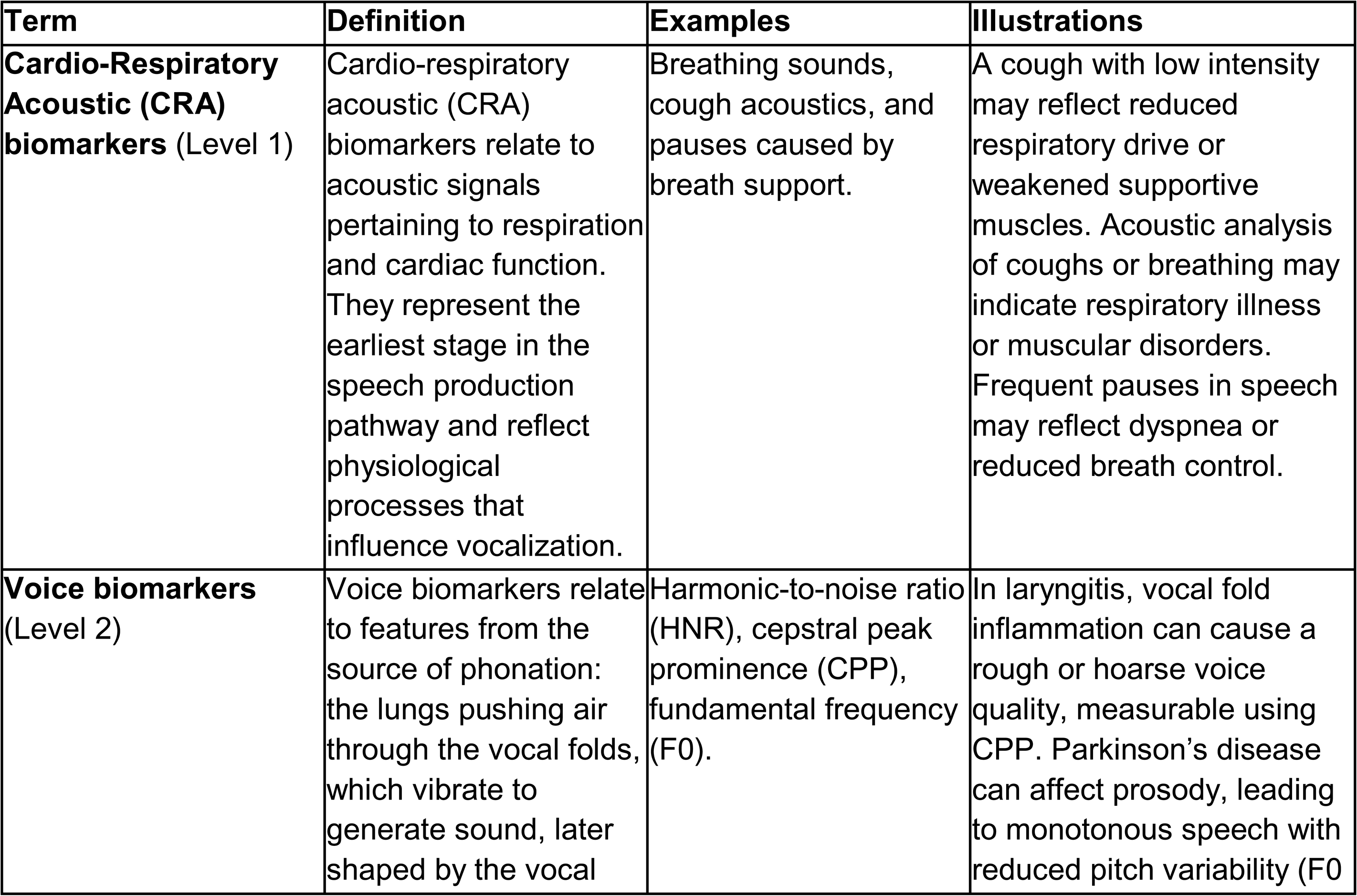
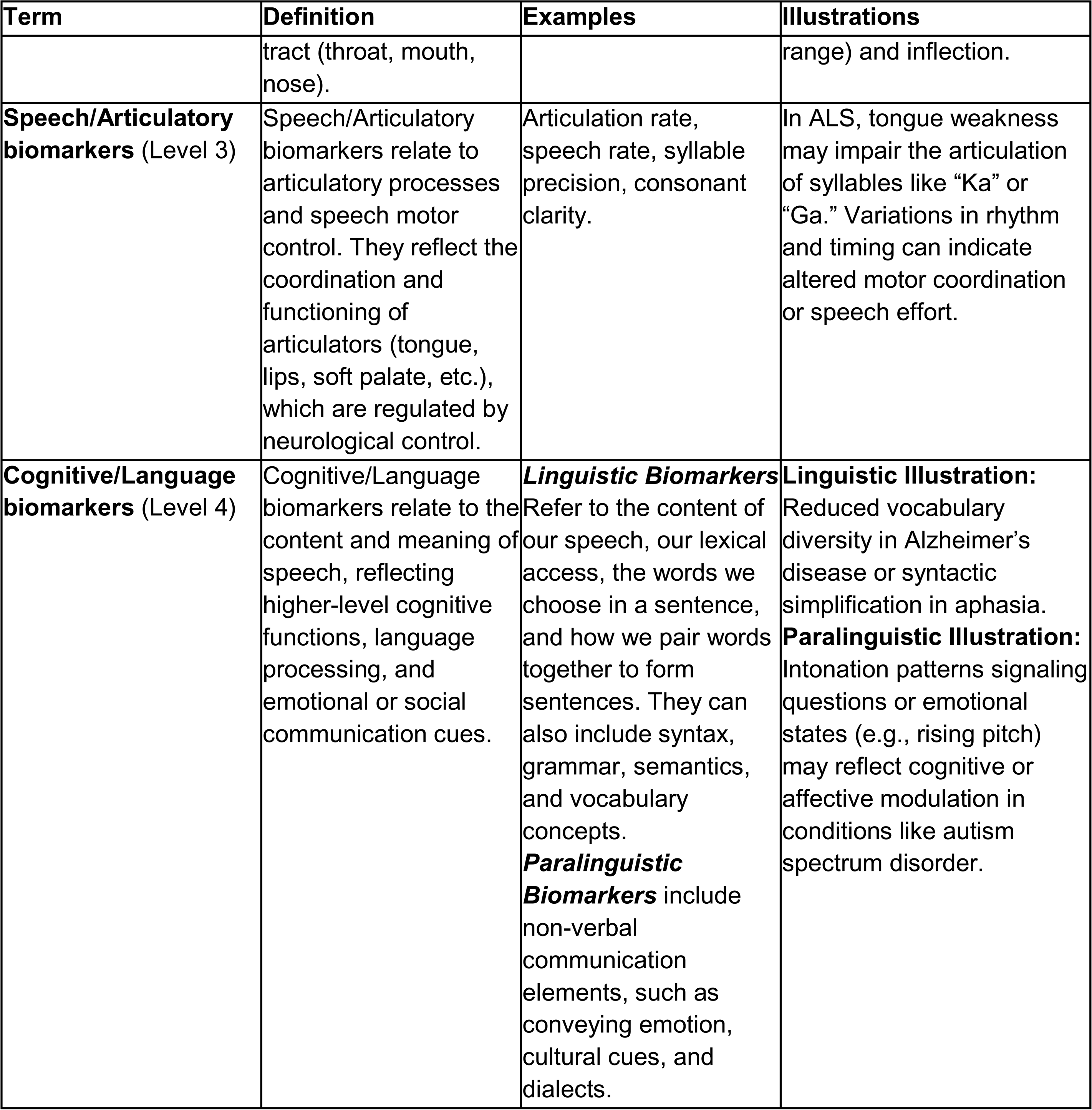
Level 1-4 Definitions.

## Discussion

This study presents the first stage of the VOCAL (Vocal Biomarker Guidelines for Ontology, Classification, Application, and Logistics) initiative, offering foundational definitions that aim to address longstanding terminological fragmentation in the field of vocal biomarkers. By introducing a layered, physiologically grounded vocabulary organized across five hierarchical levels, the framework establishes a shared reference system that facilitates interdisciplinary collaboration, supports regulatory alignment, and promotes scientific rigor. In doing so, it fills a critical gap in the biomedical and digital health landscape, where vocal biomarkers are increasingly positioned as scalable, non-invasive tools for the early detection and monitoring of diverse health conditions.

### Rethinking Voice Data and Analytics Through a Unified Lexicon

This study’s most consequential contribution is the reframing of “vocal biomarkers” not as a monolithic or interchangeable term, but as a continuum of overlapping physiological and cognitive signals. The framework intentionally avoids drawing rigid boundaries, opting instead for a layered structure that mirrors the interconnected nature of voice production, encompassing breath and muscle control, as well as language, memory, and emotion. This continuum model provides researchers and clinicians with a shared yet flexible vocabulary that can accommodate both scientific precision and real-world variability.

What makes this framework novel is its capacity to capture complexity without collapsing it. The five-level structure, from Level 0’s foundational definitions (biomarker, digital biomarker, vocal biomarker) to Level 4’s cognitive and language biomarkers, helps differentiate vocal signals not only by signal type, but also by the physiological or cognitive systems they reflect. For instance, Level 1 (cardio-respiratory acoustic biomarkers) isolates signals, such as breath pauses or cough acoustics, which are distinct from Level 2’s voice-source features, including pitch and harmonicity. Level 3 focuses on articulatory dynamics (tongue, lip, and jaw coordination), while Level 4 examines how language and social cognition are conveyed through lexical and paralinguistic cues. Importantly, this stratification does not imply mutually exclusive categories. The framework recognizes that many vocal features traverse multiple domains. Pitch, for example, may simultaneously carry phonatory, prosodic, and emotional information depending on the context. By allowing such overlaps, the model resists the oversimplification that often characterizes both academic studies and commercial applications in the voice-AI space.

This layered approach also acts as a conceptual correction to the field’s current fragmentation. Too often, research on “vocal biomarkers” has used the term without clarifying what part of the vocal system is being measured, or for what purpose. This imprecision limits interoperability between datasets, obscures reproducibility, and undermines both clinical translation and regulatory review. The proposed definitions address this gap by creating clear referents across disciplines while still allowing interpretive flexibility. Beyond classification, the framework has normative and regulatory implications. By standardizing language across a fast-evolving and commercially saturated space, it challenges the opacity of proprietary systems that often treat voice as an undifferentiated input stream. A common lexicon, co-produced by experts across laryngology, AI, neuroscience, ethics, and clinical research, fosters transparency and makes space for critical scrutiny of what is being measured, how, and why. In this way, the framework functions not only as a taxonomic scaffold but as an epistemic and ethical intervention. It realigns the field around grounded, interpretable, and shared understandings of voice and its relationship to health, an essential foundation as vocal biomarkers move from exploratory research toward clinical integration.

### Limitations

While this framework represents a significant step toward definitional coherence in vocal biomarker research, several limitations must be acknowledged. First, the process, although interdisciplinary and international, remains in its early stages. It centered primarily on researchers and professionals already embedded in consortia or academic networks. The voices of patients, regulators, and public health decision-makers from underrepresented regions were missing. Their perspectives are essential, particularly when voice data intersects with issues of trust, access, and cultural specificity. Second, the framework is intentionally definitional; it does not yet establish guidance on study design, feature extraction protocols, or validation metrics. While the definitions offer much-needed clarity on what is being measured, they do not resolve how those measurements should be standardized, interpreted, or integrated into clinical workflows. Questions regarding longitudinal stability, generalizability across languages and populations, and device dependence of vocal features remain outside the scope of this initial effort. Third, the flexible continuum approach (while a strength conceptually) may present practical challenges in operationalization. Some features can be categorized at more than one level, depending on their context. This ambiguity is intentional and reflects the non-discrete nature of vocal production, but it may complicate automated classification, regulatory labeling, or systematic reviews that require sharper boundaries. Finally, while the consensus process was rigorous and iterative, it does not constitute empirical validation. Whether these definitions improve reproducibility, regulatory acceptance, or interdisciplinary collaboration remains to be seen. The framework should thus be seen as provisional and living: a starting point for dialogue, not a final verdict.

### Contribution to Vocal Biomarkers Development and Clinical Uptake

The definitional framework proposed in this paper addresses a longstanding barrier in the field of vocal biomarkers: the lack of precise and consistent terminology. By structuring vocal biomarkers along a physiologically grounded continuum, the framework offers a common language for describing and interpreting voice-based health data. This contribution extends beyond academic precision, laying the groundwork for broader adoption, accountability, and clinical relevance.

Standardization acts as a catalyst for trust and uptake across the healthcare ecosystem. When definitions are unambiguous and widely accepted, clinicians are better equipped to interpret findings, patients are more likely to trust voice-based technologies, and regulators can more efficiently evaluate and approve them. This creates a reinforcing cycle: clarity builds trust, trust encourages adoption, and adoption drives further research, innovation, and investment. The framework also addresses a significant challenge in digital health: the opacity of proprietary systems. Open, transparent definitions reduce the “black box” problem by making it easier to understand what is being measured, how it is measured, and why. This fosters reproducibility, supports ethical governance, and enables cross-study comparisons, thereby reducing duplication and accelerating progress. For patients and clinicians alike, shared terminology enhances communication and facilitates the meaningful use of healthcare information. For third-party payers (e.g., private and public insurers) and regulators, standardization reduces uncertainty and facilitates alignment with existing biomarker approval pathways. In all cases, the framework advances the conditions necessary for responsible innovation and sustainable integration of vocal biomarkers into healthcare systems.

### Future Research

This framework offers a foundation, not an endpoint. Future research should test the practical utility of the continuum model across various use cases, examining whether it improves consistency in feature labeling, interpretability, and reproducibility in both research and clinical settings. Applying the definitions in comparative studies may clarify their impact on model transparency and cross-study compatibility. Methodological extensions are also needed, which include reporting standards aligned with the Levels, protocols for labeling multi-level features, and guidance on handling ambiguous cases. Embedding the framework into shared methodological infrastructures (such as master protocols) would support broader uptake and interoperability. Cultural, linguistic, and ethical dimensions must also be taken into account. Vocal features are shaped by more than physiology; they carry social and contextual meaning. Research must examine how definitions perform across diverse populations and communication norms, and avoid encoding bias into measurement or interpretation. Participatory approaches will be crucial in ensuring that voice-based tools are designed and deployed fairly and equitably. Finally, alignment with regulatory standards is essential. Mapping the framework to existing evidentiary categories (such as digital endpoints or AI-based diagnostics) can help establish vocal biomarkers as clinically valid and ethically deployable tools, not speculative proxies. As the field matures, this definitional groundwork should evolve alongside it, anchoring technical progress in clarity, accountability, and shared understanding.

## Conclusion

This paper presents the first structured, consensus-based definitional framework for vocal biomarkers, responding to a critical need for clarity, consistency, and interoperability in a rapidly evolving field. Developed through a rigorous, multi-stage process involving international experts across disciplines, the framework offers a shared vocabulary that reflects the physiological and cognitive dimensions of voice production. By anchoring vocal biomarkers in a layered, interpretable structure, this work provides a foundation for interdisciplinary collaboration, ethical development, and clinical translation. It helps unify fragmented research efforts, supports reproducibility, and facilitates regulatory alignment, paving the way for future guidelines, validation standards, and implementation practices. As voice continues to emerge as a rich source of health-relevant data, a common language is indispensable. This framework offers more than definitional clarity; it offers a path toward trustworthy, patient-centered, and clinically impactful voice-based health technologies.

## Data Availability

All data produced in the present work are contained in the manuscript

## Acknowledgments

We thank Claire Bortolotto for her contributions during selected meetings of the first round of review. This article/publication is based upon work from COST Action eVoiceNet (CA24128) supported by COST (European Cooperation in Science and Technology).

