## Supplementary Online Material for "Consensus-Based Definitions for Vocal Biomarkers: The International VOCAL Initiative"

### Table SOM1. Summary of the iterative process and associated modifications in the five rounds for the definitions of terms.

| **Rounds** | **Number of terms** | **Terms** | **Action plan** |
| --- | --- | --- | --- |
| **Initial draft and First Round** | 7 | - Biomarker **O** - Digital Biomarker **O** - Vocal biomarker (overarching term) **O** - Voice biomarkers **O** - Speech biomarkers **O** - Prosody biomarkers **O** - Linguistic and paralinguistic biomarkers **O** | **Panel recommendations:**   - Refine all definitions - Add *Respiratory biomarkers* and *Voice AI biomarkers* - Provide examples for each term - Include a figure |
| **Second Round** | 9 | - Biomarker **O** - Digital biomarker **O** - Vocal biomarker (overarching term) **O** - Respiratory biomarkers **O** - Voice biomarkers **O** - Speech biomarkers **O** - Prosody biomarkers **O** - Linguistic and paralinguistic biomarkers **O** - Voice AI biomarkers **O**   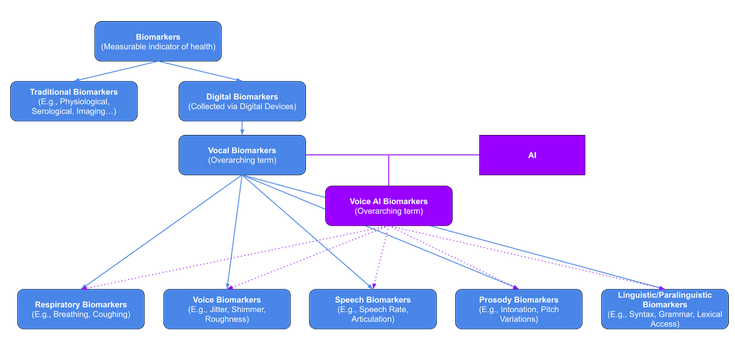 | **Panel recommendations:**   - Refine the definition of the *Biomarker* and *Digital biomarker* terms - Introduce *Acoustic biomarker* as an overarching term - Use *Vocal biomarker* as overarching term for Voice, Speech, Prosody, and Linguistic/Paralinguistic biomarkers - Replace *Respiratory biomarkers* with *Cardio-Respiratory Acoustic (CRA) biomarker* |
| **Third Round** | 10 | - Biomarker **O** - Digital biomarker **O** - Acoustic biomarker (overarching term) **O** - Vocal biomarkers (overarching term) **O** - Cardio-Respiratory Acoustic (CRA) biomarkers **O** - Voice biomarkers **O** - Speech biomarkers **O** - Prosodic biomarkers **O** - Linguistic and paralinguistic biomarkers **O** - Voice AI biomarkers **O**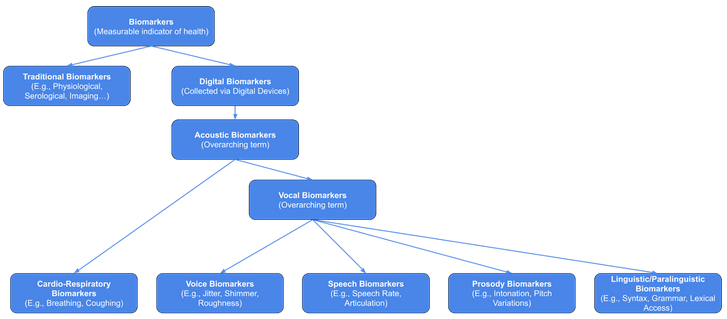 | **Panel recommendations:**   - Use *Speech biomarkers* as the overarching term for *Linguistic/Paralinguistic biomarkers* and *Prosodic biomarkers* - Refine examples |
| **Fourth Round** | 10 | - Biomarker **O** - Digital biomarker **O** - Acoustic biomarker (overarching term) **O** - Cardio-Respiratory Acoustic (CRA) biomarkers **O** - Vocal biomarkers **O** - Voice biomarkers **O** - Speech biomarkers **O** - Prosodic biomarkers **O** - Linguistic and paralinguistic biomarkers **O** - Voice AI biomarkers **O**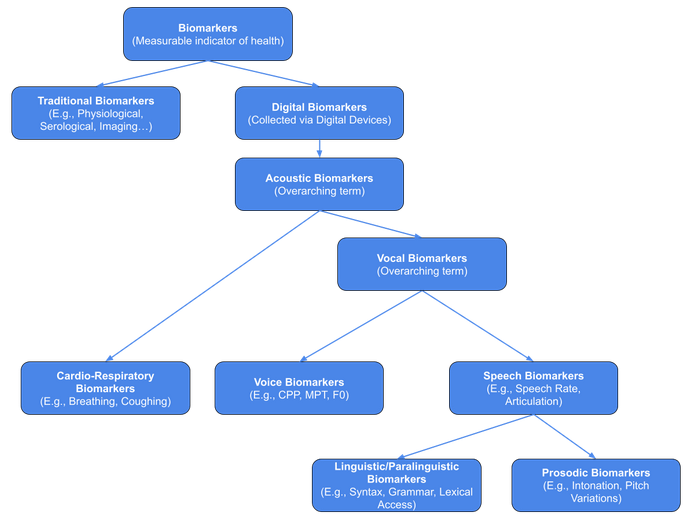 | **Panel recommendations:**   - Organize terms by level of granularity, with categorization based on the underlying physiological and cognitive systems involved in voice production - Remove *Prosodic biomarkers* as a standalone and merge them into the examples - Remove *Acoustic biomarkers* - Keep *Vocal biomarker* as the overarching term - Add the term Articulatory to *Speech/Articulatory biomarkers* - Separate *Linguistic and paralinguistic biomarkers* - Introduce *Cognitive/Language biomarkers* as an overarching term for Linguistic and Paralinguistic biomarkers - Remove *Voice AI biomarkers* |
| **Fifth Round** | 7 (+2 sub definitions) | - Level 0   - Biomarker **O**   - Digital biomarker **O**   - Vocal biomarker (overarching term) **O** - Level 1: Cardio-Respiratory Acoustic (CRA) biomarkers **O** - Level 2: Voice biomarkers **O** - Level 3: Speech/Articulatory biomarkers **O** - Level 4: Cognitive/Language biomarkers **O**   - Linguistic biomarkers **O**   - Paralinguistic biomarkers **O** | **Final structure adopted** |

**Legend**

**O**: removed / **O**: refined / **O**: added / **O**: validated

**Figure S1. Visual representation of Level 0 definitions**

*Vocal biomarkers* (blue) represent a specific subset of *digital biomarkers* (purple), which in turn are a subset of the broader category of *biomarkers* (green). The concentric circles represent increasing inclusivity, progressing from the innermost to the outermost circle. Created in [https://BioRender.com](https://biorender.com/)

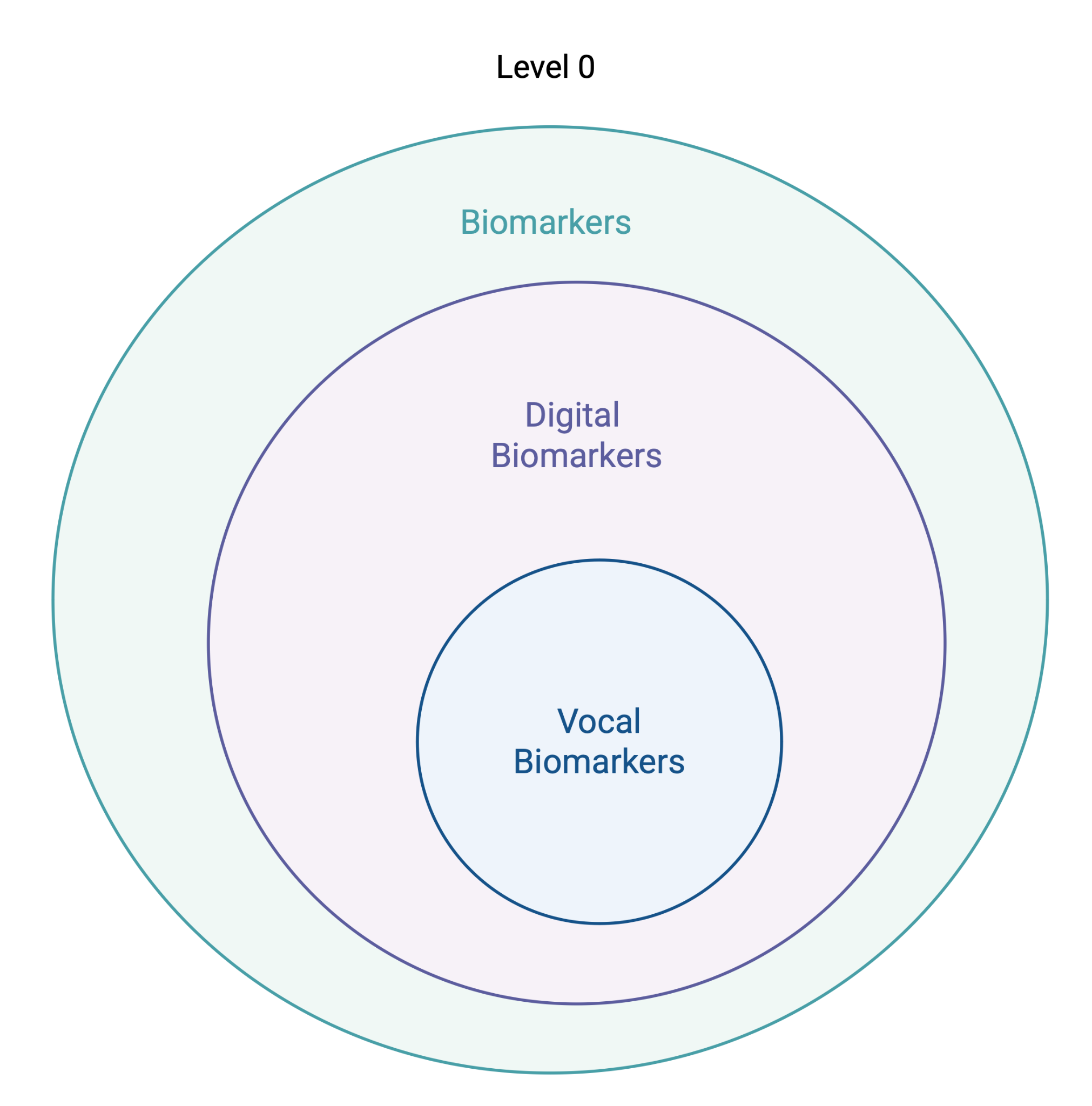
